## Supplementary file 1 for "Determinants of regulatory compliance in health and social care services: a systematic review using the Consolidated Framework for Implementation Research"

| Context | “healthcare system*” OR “health care system*” OR “care system*” OR “social care” OR ”healthcare service*” OR ”health care service*” OR ”social care service*” OR “hospital*” OR “health care setting*” OR “healthcare setting*” OR “social care setting” OR “residential facilit*” OR “care facility*” OR “nursing home*” OR “residential care” OR “long-term care” OR  “long term care” OR “disabilit*” OR “disability service” OR “care home” OR “aged care” OR “aged-care” OR “mental health service” OR “mental health centre” OR “mental health facilit*” OR “psychiatric service” OR “psychiatric centre” OR “psychiatric facilit*” OR “addiction service” OR “addiction centre” OR “addiction facilit*” OR “drug-treatment centre” OR “drug-treatment service” OR “drug-treatment facilit*” OR “drug treatment centre” OR “drug treatment service” OR “drug-treatment facilit*” OR “homecare” OR “home care” OR “domiciliary” OR “primary care” OR “community care” OR “respite care” OR “specialist care” OR “live-in care” OR “live in care” OR “homeless service*” OR “homeless shelter*” |
| --- | --- |
| Intervention | “regulation” OR “regulator*” OR “inspect*” OR “enforcement” OR “licens*” OR “certification” OR “withdrawal” OR “accredit*” |
| Mechanisms | “factor*” OR “barrier*” OR “facilitator*” OR “enabler*” OR “determinant*” OR “characteristic*” OR  “indicator*” OR “association*” OR “relationship*” OR “cause*” OR “engagement” OR “attitude*” OR “predictor*” |
| Outcome | “compliance” OR “non-compliance” OR “violat*” OR “deficienc*” OR “sanction*” OR “citation*” OR “failure*” OR “failing*” |

*Supplementary file 1 – List of search terms used for electronic database searches*
