## Supplementary file 3 for "Determinants of regulatory compliance in health and social care services: a systematic review using the Consolidated Framework for Implementation Research"

*Supplementary file 3 – Complete list of articles included in the systematic review (n=157)*

| **Title** | **Authors** | **Year** | **Country** | **Type** | **Publication name** | **Setting** |
| --- | --- | --- | --- | --- | --- | --- |
| A longitudinal study of the relationship between nurse staffing and quality of care in nursing homes | Kim, H. | 2006 | USA | Thesis | n/a | Nursing Homes |
| A national-level analysis of the relationship between nursing home satisfaction and quality | Nadash, P. et al | 2019 | USA | Journal Article | Research on Aging | Nursing Homes |
| A panel data analysis of the relationships of nursing home staffing levels and standards to regulatory deficiencies | Hongsoo, K. et al | 2009 | USA | Journal Article | The Journals of Gerontology Series B: Psychological Sciences and Social Sciences | Nursing Homes |
| A report of information technology and health deficiencies in U.S. nursing homes | Alexander, G. L.; Madsen, R. W. | 2017 | USA | Journal Article | Journal of Patient Safety | Nursing Homes |
| Abuse and neglect in nursing homes: The role of serious mental illness | Jester, D. J. et al | 2022 | USA | Journal Article | The Gerontologist | Nursing Homes |
| Administrative deficiency citations and quality of care in nursing homes | Castle, N. G.; Longest, B. B. | 2006 | USA | Journal Article | Health Services Management Research | Nursing Homes |
| Administrator turnover and quality of care in nursing homes | Castle, N. G. | 2001 | USA | Journal Article | The Gerontologist | Nursing Homes |
| An examination of special focus facility nursing homes | Castle, N. G.; Engberg, J. B. | 2010 | USA | Journal Article | The Gerontologist | Nursing Homes |
| An examination of strategic group membership and technology in the nursing home industry | Laberge, A. | 2009 | USA | Thesis | n/a | Nursing Homes |
| An exploratory study of boarding home sanctions and compliance in Washington State | Graf Schaffner, M. L. | 2011 | USA | Journal Article | Nursing Outlook | Nursing/Care Homes |
| Analysis of the relationships between nursing home administrator's types of learning and nursing home effectiveness | Loescher, S. C. | 1994 | USA | Thesis | n/a | Nursing Homes |
| Analyzing the effect of complaints, investigation of allegations, and deficiency citations on the quality of care in united states nursing homes (2007 – 2012) | Hansen, K. E. | 2015 | USA | Thesis | n/a | Nursing Homes |
| Are nursing home survey deficiencies higher in facilities with greater staff turnover | Lerner, N. B. et al | 2014 | USA | Journal Article | Journal of the American Medical Directors Association | Nursing Homes |
| Are patients with serious mental illness more likely to be admitted to nursing homes with more deficiencies in care? | Li, Y. et al | 2011 | USA | Journal Article | Medical Care | Nursing Homes |
| Association between nursing home staff turnover and infection control citations | Loomer, L. et al | 2022 | USA | Journal Article | Health Services Research | Nursing Homes |
| Building policy and service theory from nursing home inspection results: Qualitative Comparative Analysis | Bell, E. et al | 2013 | USA | Journal Article | International Journal on Disability and Human Development | Nursing Homes |
| Care deficiencies and super-organization of American nursing homes in hospital referral region | Pittman, T. | 2020 | USA | Journal Article | Frontiers in Public Health | Nursing Homes |
| Characteristics of community nursing homes serving per diem veterans, 1999 to 2002 | Johnson, C. E. et al | 2007 | USA | Journal Article | Medical Care Research and Review | Nursing Homes |
| Citations and compliance with the Nursing Home Reform Act of 1987 | Castle, N. G. | 2001 | USA | Journal Article | Journal of Health & Social Policy | Nursing Homes |
| Civil money penalty enforcement actions for quality deficiencies in nursing homes | Wang, X. et al | 2020 | USA | Journal Article | The Gerontologist | Nursing Homes |
| CMS mega-rule update and the status of pharmacy-related deficiencies in nursing homes | Wesson, K. W. | 2020 | USA | Journal Article | Journal of Applied Gerontology | Nursing Homes |
| Community presence and nursing home quality of care: the ombudsman as a complementary role | Cherry, R. L. | 1993 | USA | Journal Article | Journal of Health and Social Behavior | Nursing Homes |
| Comparing public quality ratings for accredited and nonaccredited nursing homes | Williams, S. C. et al | 2017 | USA | Journal Article | Journal of the American Medical Directors Association | Nursing Homes |
| Comparing quality of care in non-profit and for-profit nursing homes: a process perspective | Chesteen, S. et al | 2005 | USA | Journal Article | Journal of Operations Management | Nursing Homes |
| Correlation between administrator turnover and survey results | Christensen, C.; Beaver, S. | 1996 | USA | Journal Article | The Journal of Long Term Care Administration | Nursing Homes |
| Cost-effective adjustments to nursing home staffing to improve quality | Bowblis, J. R.; Roberts, A. R. | 2020 | USA | Journal Article | Medical Care Research and Review | Nursing Homes |
| Criminological theories and regulatory compliance | Makkai, T.; Braithwaite, J. | 1991 | Australia | Journal Article | Criminology | Nursing Homes |
| Cross-subsidization in nursing homes: Explaining rate differentials among payer types | Troyer, J. L. | 2002 | USA | Journal Article | Southern Economic Journal | Nursing Homes |
| Culture change and nursing home quality of care | Grabowski, D. C. et al | 2014 | USA | Journal Article | The Gerontologist | Nursing Homes |
| Deficiency citations for physical restraint use in nursing homes | Castle, N. G. | 2000 | USA | Journal Article | The Journals of Gerontology Series B: Psychological Sciences and Social Sciences | Nursing Homes |
| Determinants of non-prescription antibiotic dispensing in Chinese community pharmacies from socio-ecological and health system perspectives | Wang, X. et al | 2020 | China | Journal Article | Social Science & Medicine | Pharmacies |
| Dietary service staffing impact nutritional quality in nursing homes | Smith, K. M. et al | 2019 | USA | Journal Article | Journal of Applied Gerontology | Nursing Homes |
| Disentangling quality and safety indicator data: a longitudinal, comparative study of hand hygiene compliance and accreditation outcomes in 96 Australian hospitals | Mumford, V. et al | 2014 | Australia | Journal Article | BMJ Open | Hospitals |
| Do corporate chains affect quality of care in nursing homes? The role of corporate standardization | Kamimura, A. et al | 2007 | USA | Journal Article | Health Care Management Review | Nursing Homes |
| Do high rates of OSCAR deficiencies prompt improved nursing facility processes and outcomes? | Klopfenstein, K. et al | 2011 | USA | Journal Article | Journal of Aging & Social Policy | Nursing Homes |
| Do mandatory overtime laws improve quality? Staffing decisions and operational flexibility of nursing homes | Lu, S. F.; Lu, L. X. | 2017 | USA | Journal Article | Management Science | Nursing Homes |
| Do recipients of an association-sponsored quality award program experience better quality outcomes compared with other nursing facilities across the United States? | Castle, N. G. et al | 2018 | USA | Journal Article | Journal of Applied Gerontology | Nursing Homes |
| Does competition improve service quality? The case of nursing homes where public and private payers coexist | Lu, S. F. et al | 2021 | USA | Journal Article | Management Science | Nursing Homes |
| Does investor ownership of nursing homes compromise the quality of care? | Harrington, C. et al | 2001 | USA | Journal Article | American Journal of Public Health | Nursing Homes |
| Does litigation increase or decrease health care quality?: a national study of negligence claims against nursing homes | Stevenson, D. G. et al | 2013 | USA | Journal Article | Medical Care | Nursing Homes |
| Does the public sector outperform the nonprofit and for‐profit sectors? Evidence from a national panel study on nursing home quality and access | Amirkhanyan, A. A. et al | 2008 | USA | Journal Article | Journal of Policy Analysis and Management: The Journal of the Association for Public Policy Analysis and Management | Nursing Homes |
| Effects of Medicare payment changes on nursing home staffing and deficiencies | Konetzka, R. T. et al | 2004 | USA | Journal Article | Health Services Research | Nursing Homes |
| Effects of nursing practice environments on quality outcomes in nursing homes | Flynn, L. et al | 2010 | USA | Journal Article | Journal of the American Geriatrics Society | Nursing Homes |
| Effects of state minimum staffing standards on nursing home staffing and quality of care | Park, J.; Stearns, S. C. | 2009 | USA | Journal Article | Health Services Research | Nursing Homes |
| Elderly Hispanics more likely to reside in poor-quality nursing homes | Fennell, M. L. et al | 2010 | USA | Journal Article | Health Affairs | Nursing Homes |
| Excess demand, the percentage of Medicaid patients, and the quality of nursing home care | Nyman, J. A. | 1988 | USA | Journal Article | Journal of Human Resources | Nursing Homes |
| Exploring the challenges non-clinical departments encounter during eden alternative implementation | George, K. | 2019 | USA | Thesis | n/a | Nursing Homes |
| Exploring the influence of the regulatory survey process on nursing home administrator job satisfaction and job seeking | Holecek, T. et al | 2010 | USA | Journal Article | Journal of Applied Gerontology | Nursing Homes |
| Factors affecting compliance with national accreditation essential safety standards in the Kingdom of Saudi Arabia | Althumairi, A. et al | 2022 | Saudi Arabia | Journal Article | Scientific Reports | Hospitals |
| Factors that influence an administrator's perception of what they consider important in job knowledge and how those perceptions impact quality of care | Fabbri, M. A. | 2006 | USA | Thesis | n/a | Nursing Homes |
| Failing the metric but saving lives: The protocolization of sepsis treatment through quality measurement | Winslow, R. | 2020 | USA | Journal Article | Social Science & Medicine | Hospitals |
| Government regulation and the quality of healthcare - Evidence from minimum staffing legislation for nursing homes | Matsudaira, J. D. | 2014 | USA | Journal Article | Journal of Human Resources | Nursing Homes |
| Hand hygiene deficiency citations in nursing homes | Castle, N. G. et al | 2014 | USA | Journal Article | Journal of Applied Gerontology | Nursing Homes |
| Hidden owners, hidden profits, and poor nursing home care | Harrington, C. et al | 2015 | USA | Journal Article | International Journal of Health Services | Nursing Homes |
| High-performing and low-performing nursing homes: a view from complexity science | Forbes-Thompson, S. et al | 2007 | USA | Journal Article | Health Care Management Review | Nursing Homes |
| Hospital characteristics associated with penalties in the centers for Medicare & Medicaid Services Hospital-Acquired Condition Reduction Program | Rajaram, R. et al | 2015 | USA | Journal Article | Journal of the American Medical Association | Hospitals |
| Impact of voluntary accreditation on deficiency citations in U.S. nursing homes | Wagner, L. M. et al | 2012 | USA | Journal Article | The Gerontologist | Nursing Homes |
| Implementation of an emergency power rule: Compliance of Florida nursing homes and assisted living facilities | June J. W. et al | 2022 | USA | Journal Article | Disaster Medicine and Public Health Preparedness | Nursing Home/Assisted-Living Facilities |
| Infection control citations in nursing homes: Compliance and geographic variability | Jester, D. J. et al | 2021 | USA | Journal Article | Journal of the American Medical Directors Association | Nursing Homes |
| Infection prevention and control programs in US nursing homes: results of a national survey | Herzig, C. T. et al | 2016 | USA | Journal Article | Journal of the American Medical Directors Association | Nursing Homes |
| Influence of administrator and facility characteristics on nursing home performance | Singh, D. A. | 1994 | USA | Thesis | n/a | Nursing Homes |
| Institutional form and the nursing home industry: Ownership effects on costs and quality | Holmes, J. S. | 1992 | USA | Thesis | n/a | Nursing Homes |
| Intended and unintended consequences of minimum staffing standards for nursing homes | Chen, M. M.; Grabowski, D. C. | 2015 | USA | Journal Article | Health Economics | Nursing Homes |
| Iowa nursing home characteristics associated with reported abuse | Jogerst, G. J. et al | 2006 | USA | Journal Article | Journal of the American Medical Directors Association | Nursing Homes |
| Is nurse aide retention associated with nursing home quality? | Kennedy, K. A. | 2021 | USA | Thesis | n/a | Nursing Homes |
| Is the quality of nursing homes countercyclical? Evidence from 2001 through 2015 | Huang, S. S.; Bowblis, J. R. | 2019 | USA | Journal Article | The Gerontologist | Nursing Homes |
| Joint commission accreditation and quality measures in US nursing homes | Wagner, L. M. et al | 2012 | USA | Journal Article | Policy, Politics, & Nursing Practice | Nursing Homes |
| Long-term care standards: enforcement and compliance | Christianson, J. B. | 1979 | USA | Journal Article | Journal of Health Politics, Policy and Law | Nursing Homes |
| Medicare's prospective payment system for skilled nursing facilities: Effects on staffing and quality of care | White, C. | 2005 | USA | Journal Article | INQUIRY: The Journal of Health Care Organization, Provision, and Financing | Nursing Homes |
| Mental health care deficiency citations in nursing homes and caregiver staffing | Castle, N. G.; Myers, S. | 2006 | USA | Journal Article | Administration and Policy in Mental Health and Mental Health Services Research | Nursing Homes |
| Nurse aide retention in nursing homes | Castle, N. G. et al | 2020 | USA | Journal Article | The Gerontologist | Nursing Homes |
| Nurse staffing and deficiencies in the largest for-profit nursing home chains and chains owned by private equity companies | Harrington, C. et al | 2012 | USA | Journal Article | Health Services Research | Nursing Homes |
| Nurse staffing and deficiency of care for inappropriate psychotropic medication use in nursing home residents with dementia | Yoon, J. M. et al | 2022 | USA | Journal Article | Journal of Nursing Scholarship | Nursing Homes |
| Nurse staffing and quality of care in nursing facilities | Johnson-Pawlson, J.;  Infeld, D. L. | 1996 | USA | Journal Article | Journal of Gerontological Nursing | Nursing Homes |
| Nurse staffing levels and quality of care in Northeastern Pennsylvania nursing homes | Akinci, F.; Krolikowski, D. | 2005 | USA | Journal Article | Applied Nursing Research | Nursing Homes |
| Nursing home administrator turnover and quality of care: A quantitative study | Harvey, D. | 2014 | USA | Thesis | n/a | Nursing Homes |
| Nursing home deficiencies: An exploratory study of interstate variations in regulatory activity | Kelly, C. M. et al | 2008 | USA | Journal Article | Journal of Aging & Social Policy | Nursing Homes |
| Nursing home deficiency citations for abuse | Castle, N. G. | 2011 | USA | Journal Article | Journal of Applied Gerontology | Nursing Homes |
| Nursing home deficiency citations for infection control | Castle, N. G. et al | 2011 | USA | Journal Article | American Journal of Infection Control | Nursing Homes |
| Nursing home deficiency citations for medication use | Castle, N. G.; Engberg, J. B. | 2007 | USA | Journal Article | Journal of Applied Gerontology | Nursing Homes |
| Nursing home deficiency citations for physical restraints and restrictive side rails | Wagner, L. M. et al | 2008 | USA | Journal Article | Western Journal of Nursing Research | Nursing Homes |
| Nursing home deficiency citations for safety | Castle, N. G. et al | 2010 | USA | Journal Article | Journal of Aging & Social Policy | Nursing Homes |
| Nursing home director of nursing leadership style and director of nursing-sensitive survey deficiencies | McKinney, S. H. et al | 2016 | USA | Journal Article | Health Care Management Review | Nursing Homes |
| Nursing home environment and organizational performance: Association with deficiency citations | Temkin-Greener, H. et al | 2010 | USA | Journal Article | Medical Care | Nursing Homes |
| Nursing home infection control program characteristics, CMS citations, and implementation of antibiotic stewardship policies: A national study | Stone, P. W. et al | 2018 | USA | Journal Article | INQUIRY: The Journal of Health Care Organization, Provision, and Financing | Nursing Homes |
| Nursing home profit margins and citations for infection prevention and control | Sharma, H.; Xu, L. | 2021 | USA | Journal Article | Journal of the American Medical Directors Association | Nursing Homes |
| Nursing home quality and financial performance: Does the racial composition of residents matter? | Chisholm, L. et al | 2013 | USA | Journal Article | Health Services Research | Nursing Homes |
| Nursing home quality and financial performance: Is there a business case for quality? | Weech-Maldonado, R. et al | 2019 | USA | Journal Article | INQUIRY: The Journal of Health Care Organization, Provision, and Financing | Nursing Homes |
| Nursing home safety: does financial performance matter? | Oetjen, R. M. et al | 2011 | USA | Journal Article | Journal of Health Care Finance | Nursing Homes |
| Nursing home staffing and its relationship to deficiencies | Harrington, C. et al | 2000 | USA | Journal Article | The Journals of Gerontology Series B: Psychological Sciences and Social Sciences | Nursing Homes |
| Nursing home survey deficiencies for physical restraint use | Graber, D. R.; Sloane, P. D. | 1995 | USA | Journal Article | Medical Care | Nursing Homes |
| Nursing home work environment characteristics: Associated outcomes in psychosocial care | Bonifas, R. P. | 2008 | USA | Journal Article | Health Care Financing Review | Nursing Homes |
| Nursing homes in states with infection control training or infection reporting have reduced infection control deficiency citations | Cohen, C. C. et al | 2015 | USA | Journal Article | Infection Control & Hospital Epidemiology | Nursing Homes |
| Nursing homes with persistent deficiency citations for physical restraint use | Castle, N. G. | 2002 | USA | Journal Article | Medical Care | Nursing Homes |
| Nursing staff availability and other facility characteristics in relation to assisted living care deficiencies | Trinkoff, A. M. et al | 2019 | USA | Journal Article | Journal of Nursing Regulation | Assisted-living facilities |
| Organizational and environmental effects on voluntary and involuntary turnover | Donoghue, C.; Castle, N. G. | 2007 | USA | Journal Article | Health Care Management Review | Nursing Homes |
| Organizational correlates of the risk-adjusted pressure ulcer prevalence and subsequent survey deficiency citation in California nursing homes | Dellefield, M. E. | 2006 | USA | Journal Article | Research in Nursing & Health | Nursing Homes |
| Outcomes of health information technology utilization in nursing homes: Do implementation processes matter? | Hamann, D. J.; Bezboruah, K. C. | 2020 | USA | Journal Article | Health Informatics Journal | Nursing Homes |
| Ownership conversions and nursing home performance | Grabowski, D. C.; Stevenson, D. G. | 2008 | USA | Journal Article | Health Services Research | Nursing Homes |
| Performative compliance and the state–corporate structuring of neglect in a residential care home for older people | Greener, J. | 2019 | UK | Journal Article | Critical Criminology | Nursing Homes |
| Physician compliance with quality and patient safety regulations: The role of perceived enforcement approaches and commitment | Weske, U. et al | 2019 | Netherlands | Journal Article | Health Services Management Research | Hospitals |
| Predictors of quality in rural nursing homes using standard and novel methods | Towsley, G. L. et al | 2013 | USA | Journal Article | Research in Gerontological Nursing | Nursing Homes |
| Predictors of quality of care in California nursing homes | Dellefield, M. E. | 1999 | USA | Thesis | n/a | Nursing Homes |
| Providing outcomes information to nursing homes: can it improve quality of care? | Castle, N. G. | 2003 | USA | Journal Article | The Gerontologist | Nursing Homes |
| Public, private, nonprofit ownership and the response to asymmetric information: The case of nursing homes | Weisbrod, B. A.; Schlesinger, M. | 1986 | USA | Book Section | The Economics of Nonprofit Institutions: Studies in Structure and Policy | Nursing Homes |
| Quality care and financial viability of rural nursing homes | Towsley, G. L. | 2007 | USA | Thesis | n/a | Nursing Homes |
| Quality concerns in nursing homes that serve large proportions of residents with serious mental illness | Jester, D. J. et al | 2020 | USA | Journal Article | The Gerontologist | Nursing Homes |
| Quality failures in residential aged care in Australia: The relationship between structural factors and regulation imposed sanctions | Baldwin, R. et al | 2015 | Australia | Journal Article | Australasian Journal on Ageing | Nursing Homes |
| Quality indicator survey versus traditional survey in New York State: a comparison of results from annual nursing home surveys | Delaney, C. M. et al | 2018 | USA | Journal Article | Journal of Aging & Social Policy | Nursing Homes |
| Quality of care and California nursing homes: The effects of staffing, organizational, resident and market characteristics | Collier, E. J. | 2008 | USA | Thesis | n/a | Nursing Homes |
| Quality of care in nursing homes: An analysis of relationships among profit, quality, and ownership | O'Neill, C. et al | 2003 | USA | Journal Article | Medical Care | Nursing Homes |
| Quality of care: Impact of nursing home characteristics | Lee, H. Y. | 2009 | USA | Thesis | n/a | Nursing Homes |
| Quality procedures and complaints: nursing homes in Portugal | Gil, A. P. | 2019 | Portugal | Journal Article | The Journal of Adult Protection | Nursing Homes |
| Reducing antipsychotic medication use in nursing homes: A qualitative study of nursing staff perceptions | Simmons, S. F. et al | 2018 | USA | Journal Article | The Gerontologist | Nursing Homes |
| Reforming nursing home quality regulation: Impact on cited deficiencies and nursing home outcomes | Spector, W. D.; Drugovich, M. L. | 1989 | USA | Journal Article | Medical Care | Nursing Homes |
| Registered nurse staffing and OBRA deficiencies in Nevada nursing facilities | Moseley, C. B.; Jones, L. | 2003 | USA | Journal Article | Journal of Gerontological Nursing | Nursing Homes |
| Registered nurse staffing mix and quality of care in nursing homes: A longitudinal analysis | Kim, H. et al | 2009 | USA | Journal Article | The Gerontologist | Nursing Homes |
| Regulatory styles, motivational postures and nursing home compliance | Braithwaite, V. et al | 1994 | Australia | Journal Article | Law & Policy | Nursing Homes |
| Reintegrative shaming and compliance with regulatory standards | Makkai, T.; Braithwaite, J. | 1994 | Australia | Journal Article | Criminology | Nursing Homes |
| Relationship between nurse staffing and quality of care in Louisiana nursing homes | Kercado, V. | 2016 | USA | Thesis | n/a | Nursing Homes |
| Relationship between quality of care and negligence litigation in nursing homes | Studdert, D. M. et al | 2011 | USA | Journal Article | New England Journal of Medicine | Nursing Homes |
| Relationship of facility characteristics and presence of an Ombudsman to Missouri long-term care facility state inspection report results | Cox, C. C. et al | 2011 | USA | Journal Article | Journal of Elder Abuse & Neglect | Nursing Homes |
| Relationship of ownership and size to quality in Wisconsin nursing homes | Riportella-Muller, R.; Slesinger, D. P. | 1982 | USA | Journal Article | The Gerontologist | Nursing Homes |
| Reliability of the nursing home survey process: A simultaneous survey approach | Lee, R. H. et al | 2006 | USA | Journal Article | The Gerontologist | Nursing Homes |
| Revisiting the relationship between nurse staffing and quality of care in nursing homes: An instrumental variables approach | Lin, H. | 2014 | USa | Journal Article | Journal of Health Economics | Nursing Homes |
| Serious mental illness and nursing home quality of care | Rahman, M. et al | 2013 | USA | Journal Article | Health Services Research | Nursing Homes |
| Staff turnover and quality of care in nursing homes | Castle, N. G.; Engberg, J. B. | 2005 | USA | Journal Article | Medical Care | Nursing Homes |
| Staffing-related deficiency citations in nursing homes | McDonald, S. M. et al | 2013 | USA | Journal Article | Journal of Aging & Social Policy | Nursing Homes |
| State nursing home enforcement systems | Harrington, C. et al | 2004 | USA | Journal Article | Journal of Health Politics, Policy and Law | Nursing Homes |
| Testing an expected utility model of corporate deterrence | Braithwaite, J; Makkai, T. | 1991 | Australia | Journal Article | Law & Society Rev. | Nursing Homes |
| The admission of blacks to high-deficiency nursing homes | Grabowski, D. C. | 2004 | USA | Journal Article | Medical Care | Nursing Homes |
| The association between hospital characteristics and emergency medical treatment and Labor Act citation events | Terp, S. et al | 2020 | USA | Journal Article | Medical Care | Hospitals |
| The association between staff retention and English care home quality | Allan, S.; Vadean, F. | 2021 | UK | Journal Article | Journal of Aging & Social Policy | Nursing/Care Homes |
| The contribution of electronic health records to risk management through accreditation of residential aged care homes in Australia | Yu, P. et al | 2020 | Australia | Journal Article | BMC Medical Informatics and Decision Making | Nursing Homes |
| The effects of ownership and ownership change on nursing home industry costs | Holmes, J. S. | 1996 | USA | Journal Article | Health Services Research | Nursing Homes |
| The impact of Electronic Health Records on risk management of information systems in Australian residential aged care homes | Jiang, T. et al | 2016 | Australia | Journal Article | Journal of Medical Systems | Nursing Homes |
| The impact of nurse turnover on quality of care and mortality in nursing homes: Evidence from the great recession | Antwi, Y. A.; Bowblis J. R. | 2018 | USA | Journal Article | American Journal of Health Economics | Nursing Homes |
| The influence of consistent assignment on nursing home deficiency citations | Castle, N. G. | 2011 | USA | Journal Article | The Gerontologist | Nursing Homes |
| The influence of nurse staffing levels on quality of care in nursing homes | Hyer, K. et al | 2011 | USA | Journal Article | The Gerontologist | Nursing Homes |
| The influence of nursing home characteristics and task environment on complaints and survey performance | Graber, D. R. | 1993 | USA | Thesis | n/a | Nursing Homes |
| The influence of registered nurse staffing on the quality of nursing home care | Munroe, D. J. | 1990 | USA | Journal Article | Research in Nursing & Health | Nursing Homes |
| The quality indicator survey: Background, implementation, and widespread change | Lin, M. K.; Kramer, A. M. | 2013 | USA | Journal Article | Journal of Aging & Social Policy | Nursing Homes |
| The quality of care in residential care facilities for the elderly | Flores, C. | 2007 | USA | Thesis | n/a | Nursing Home/Assisted-Living Facilities |
| The regulation and enforcement of federal nursing home standards, 1991-1997 | Harrington, C; Carrillo, H. | 1999 | USA | Journal Article | Medical Care Research and Review | Nursing Homes |
| The regulation of US nursing homes: An examination of state and federal tools and their effect on providers' performance | Hawks, B. A. | 2018 | USA | Thesis | n/a | Nursing Homes |
| The relationship between nursing staff levels, skill mix, and deficiencies in Maryland nursing homes | Lerner, N. B. | 2013 | USA | Journal Article | The Health Care Manager | Nursing Homes |
| The relationship between volunteer long-term care ombudsmen and regulatory nursing home actions | Nelson, H. W. et al | 1995 | USA | Journal Article | The Gerontologist | Nursing Homes |
| The rise of human service chains: antecedents to acquisitions and their effects on the quality of care in US nursing homes | Banaszak-Holl, J. et al | 2002 | USA | Journal Article | Managerial and Decision Economics | Nursing Homes |
| The role of sanctions in Australia's residential aged care quality assurance system | Ellis, J. M.; Howe, A. | 2010 | Australia | Journal Article | International Journal for Quality in Health Care | Nursing Homes |
| The role of the not-for-profit firm in the mixed industry: Three empirical analyses of the long-term care and hospital industries | Ballou, J. P. | 2000 | USA | Thesis | n/a | Nursing Homes |
| The social worker in interdisciplinary care planning | Vongxaiburana, E. et al | 2011 | USA | Journal Article | Clinical Gerontologist | Nursing Homes |
| The use of contract licensed nursing staff in U.S. nursing homes | Bourbonniere, M. et al | 2006 | USA | Journal Article | Medical Care Research and Review | Nursing Homes |
| Unannounced versus announced hospital surveys: a nationwide cluster-randomized controlled trial | Ehlers, L. H. et al | 2017 | Denmark | Journal Article | International Journal for Quality in Health Care | Hospitals |
| Using deficiency data to measure quality in assisted living communities: A Florida statewide study | June J. W. et al | 2020 | USA | Journal Article | Journal of Aging & Social Policy | Assisted-living facilities |
| Voluntary and involuntary nursing home staff turnover | Donoghue, C.; Castle, N. G. | 2006 | USA | Journal Article | Research on Aging | Nursing Homes |
| Who are the innovators? Nursing homes implementing culture change | Grabowski, D. C. et al | 2014 | USA | Journal Article | The Gerontologist | Nursing Homes |
| Why are some care homes better than others? An empirical study of the factors associated with quality of care for older people in residential homes in Surrey, England | Gage, H. et al | 2009 | UK | Journal Article | Health & Social Care in the Community | Nursing Homes |
