## Supplementary file 6 for "Determinants of regulatory compliance in health and social care services: a systematic review using the Consolidated Framework for Implementation Research"

*Supplementary file 6 – Author reflections on using the Consolidated Framework for Implementation Research*

Authors using the CFIR are encouraged to reflect on three questions: (1) coherence of terminology, (2) whether use of the CFIR promotes comparison across studies, and (3) whether use stimulates new theoretical development.^(1)^

To address question 1, we found the terminology of the CFIR to be broadly coherent despite overlap across some constructs. For example, network-related determinants could be coded to either the ‘cosmopolitanism’ or ‘networks & communications’ constructs. Similarly, we encountered determinants that could have been coded to both the ‘culture’ and ‘knowledge and beliefs about the innovation’ constructs.

We also found a significant number of determinants for which there was no appropriate construct. We coded these as ‘other’ within the most appropriate domain. Any decision to code a determinant as ‘other’ was principally done because the studies did not make any explicit link with the implementation of regulations. For example, some determinants related to costs or finances were coded as ‘other’ because they could not be linked to costs associated with implementing regulations. This is not a criticism of the CFIR but more a feature of the literature in this review.

We would also concur with the previously identified gap in the CFIR around implementation sustainability. The implementation of regulations and the ongoing process by which compliance is measured means that the process does not simply cease at the point of ‘reflecting and evaluating’. As a final point to address question 1, our review found many determinants that related to people’s medical conditions or care needs. We could not code such determinants to this construct because it was limited to how well an organisation *accurately knew and prioritised* the needs of these service users, as opposed to those people’s actual or potential needs. This was also an observation made by authors of another study using the CFIR.^(2)^

On question 2, cross-comparison was not a possibility due to the nature of our study and the lack of any similar studies in this field using the CFIR. However, there may be merit in utilising the CFIR across jurisdictions or across distinct service types to compare findings on determinants of regulatory compliance.

On question 3, the process of deciding on the coding of individual determinants raised some useful discussion among the authors. We ultimately decided on coding a large number of determinants to ‘other’ in each of the five domains. These were primarily related to externalities, determinants over which an organisation had little or no control, or were entirely unrelated to implementing regulations, but may have had a bearing on that organisation’s ability to successfully implement regulations (e.g. staffing, population density, political climate). This prompted discussions among the authors as to whether there was a suitable place in the CFIR to address phenomena that are extraneous to organisations and also to the implementation process, but nevertheless influence the success of an innovation. Indeed, a similar observation was made in another study that used the CFIR.^(3)^
